# Processes for creating an interprofessional mental health identity among pre-registration healthcare students: A scoping review protocol

**DOI:** 10.1101/2025.04.30.25326712

**Authors:** Melissa Owens, Charl Filies Gerard, Molly Crosland, Claire Carswell

## Abstract

**Background:** Developing an interprofessional mental health identity among pre-registration (licensure) healthcare students is critical for promoting effective interprofessional collaboration in mental health care. An interprofessional mental health identity refers to the shared beliefs, values and attitudes that will enable healthcare students/professionals to work together effectively in addressing mental health issues. An interprofessional socialisation framework can assist students and healthcare professionals develop a dual identity to foster interprofessional collaborative person-centred practice. However, the processes and strategies for facilitating this amongst pre-registration healthcare students are not clearly defined or understood. This scoping review aims to map the available literature on how interprofessional mental health identity is created for pre-registration healthcare students.

**Methods:** We will follow the five stages of Arksey and O’Malley’s Framework and identify our search terms using the ‘population’, ‘concept’ and ‘context’ (PCC) criteria. We will search CINAHL, Medline, British Nursing Database, PsychInfo, Cochrane Library and AMED from 1990 to May 2024. Retrieved records will be managed in Covidence and screened independently by two reviewers. Data extraction forms will be developed to capture relevant data related to the review aim. The forms will be piloted and the data extraction process completed by two independent reviewers. Narrative synthesis will be used to provide a descriptive overview of the included articles.

**Discussion:** Little is known regarding the process by which pre-registration healthcare students develop an interprofessional, mental health identity. Therefore, this scoping review will draw together existing research to create a conceptual model for the process of developing an interprofessional mental health identity.

## Introduction

Mental health is a neglected but crucial aspect of overall well-being, and its importance in healthcare has been increasingly recognised in recent years. Votruba and Thornicroft (2016) [1] highlight the growing importance of mental health in healthcare. In September 2015, mental health was included in the United Nations (UN) Sustainable Development Goals (SDGs). The UN, therefore acknowledged for the next 15 years, the burden of disease of mental illness as a growing priority for global development. Nevertheless, mental health orders remain at an all-time high and access to effective mental health services continues to be a significant challenge globally for the population [2]. The need for professionals to work together collaboratively to address this challenge has long since been established [3], with interprofessional collaboration specifically identified as a key action for improving the quality and efficiency of mental health services [4]. For the health workforce to collaboratively address health challenges, interprofessional education (IPE) is needed to lay down a foundation. IPE aims to prepare pre-registration healthcare students from different professions to work collaboratively and develop a shared understanding of their roles and responsibilities in providing patient-centred care [2].

We believe, therefore, that developing an interprofessional mental health identity among pre- registration healthcare students is critical for promoting effective interprofessional collaboration in mental health care. An interprofessional identity refers to the shared beliefs, values and attitudes that will enable healthcare students/professionals to work together effectively [5]. However, the processes and strategies for fostering an interprofessional mental health identity among pre- registration healthcare students are not clearly defined or understood.

The concept of an interprofessional identity first emerged in the 1990s and, although no definitive definition exists, is helpfully described by Khalili et al. as a ‘sense of belonging to own profession and interprofessional community’[5] and is considered to be made up of three elements: a sense of belonging, of commitment and of shared beliefs [6] . It is essential to contemporary practice to enhance interprofessional working practices, although particular challenges are recognised in achieving this at pre-registration level. In particular, power differentials can reduce confidence in speaking up and ultimately affect patient safety [7]. Discussions are continuing regarding how best to facilitate the development of an interprofessional identity, with challenges such as power dynamics recognised as a factor that can impede it [7]. Sociological literature, such as Witz [8] and Freidson [9], has been used to consider how identity is formed within professional groups, with identity theory, as well as social identity theory, used to explore and explain interprofessional identity [7, 10].

However, pre-registration students face particular challenges in finding a mental health field identity due to the generic nature of most professional programmes. Globally, for example, nursing students will complete a generic programme of study and then specialise in a field of practice after registration. However, in the UK, nursing students choose their field of practice at the point of application, meaning the professional identity for mental health field students could be formed early in their professional careers.

However, a significant challenge they face is the genericism of the nursing programmes within the UK. The Professional Standards for Nursing [11] require students from all fields of nursing to be competent in the same proficiencies which can lead to a sense of hierarchy between fields of nursing, with mental health nursing seen as being of lower status than that of the adult field nurse [12]. Pre-registration students have a strong desire to ‘belong’ to their own profession before and in the early stages of their professional journey [13, 14], which can lead to resentment of others [15].

Having a strong, professional identity is considered an essential prerequisite to developing a strong, interprofessional, also known as a dual, identity [5]. However, it can equally have an opposite effect if the uni-professional identity feels threatened [15] and thus makes this review important.

### Aim and Objectives

The aim of this review is to map the available literature on how interprofessional mental health identity has been created for pre-registration healthcare students and includes two overarching objectives:

⍰ Outline the range and nature of literature published on interprofessional mental health identity for pre-registration healthcare students
⍰ IDescribe the processes and strategies used to foster an interprofessional mental health identity in pre-registration healthcare students

## Materials and Methods

A scoping review will be conducted to address the breadth of the research question and map the available literature on the subject. This scoping review will follow the five stages of Arksey and O’Malley’s (2005) Framework [16]. The protocol has been registered prospectively on Open Science Framework Registries (DOI: https://doi.org/10.17605/OSF.IO/TJ9E7) prior to screening records. At the time of writing this protocol, data extraction has not yet started.

### Stage One: Identification of the Research Question

To focus on mental health identity in pre-registration health care students and ascertain if an interprofessional mental health identity has been defined and, if so, the processes by which it is created.

### Stage Two: Identify Relevant Studies

#### Eligibility Criteria

Inclusion and exclusion criteria were developed according to the PCC mnemonic (Population, Concept, Context).

We will include:

⍰ Articles that focus on a population of pre-registration healthcare students or involve participants who are pre-registration healthcare students. In this review, this is defined as anyone undertaking a degree in higher education that would make them eligible for registration with a professional regulatory body, enabling them to work in the area of mental health. This would include, but is not limited to, nurses, medical doctors, occupational therapists, pharmacists, other allied healthcare professionals, clinical psychologists and social workers.
⍰ Articles which describe interprofessional mental health identity among pre-registration healthcare students.
⍰ Articles which describe the processes of creating an interprofessional mental health identity.
⍰ Primary empirical research articles (using any methodology), opinion pieces, theoretical articles, news articles, governmental or charity reports, white papers, editorial letters, commentary, and other grey literature.
⍰ Articles published in English. I] Articles published after 1990.

We will exclude:

⍰ Articles which focus exclusively on post-registration healthcare professionals.
⍰ Articles which focus exclusively on healthcare workers or students enrolled on a course or degree which does not make them eligible for registration with a professional regulatory body, or who will not be subjected to statutory regulation before May 2024. For example, physician associates, or healthcare support workers.
⍰ Articles which describe the processes of creating an interprofessional identity outside the context of mental health.
⍰ Articles published in a language other than English, due to a lack of resources for translation services.
⍰ Articles published before 1990, due to the changes in education and regulation of healthcare workers prior to 1990.

#### Information sources

We will search the following electronic databases: CINAHL, Medline, British Nursing Database, PsychInfo, Cochrane Library, AMED from 1990 to May 2024 using search terms relevant to the population, concept and context of the review. These search terms were developed by identifying the relevant population, concept, context, and key terms that would capture these aspects of the review question. We also identified previous reviews in relevant areas to identify important terms and MeSH headings for inclusion in the search strings. We refined the search strings through consultation with a subject librarian and piloting the search strings on Medline and CINAHL. An exemplar search strategy for Medline is available in Appendix 1. We will identify grey literature through searches of EThOs and ProQuest to find relevant dissertations and theses and search for evaluation, policy documents and white papers from relevant organisations, such as Interprofessional.Global (IP.Global), Interprofessional Research.Global (IPR.Global), The Centre for the Advancement for Interprofessional Education (CAIPE), The Africa Interprofessional Education Network (AfrIPEN), The Asia Pacific Interprofessional Education and Collaboration Network (APIPEC), the Arab Network for Interprofessional Collaboration (ANIC), The Australian Interprofessional Practice and Education Network (AIPPEN), The National Center for Interprofessional Practice and Education (AIHC), The Noric Interprofessional Network (NIPNET), The Regional Network for Interprofessional Education in the Americas (REIP), The National Academies of Practice (NAP), The Dutch-speaking Network for Interprofessional Collaboration (IPINN), The Indian Interprofessional Education Network (IndIPEN) and, IP-Health. Check re Canada

We will hand-search specialist journals that publish research on interprofessional working in health and social care, such as the Journal of Interprofessional Care, and consult with relevant experts in the field to ensure that all relevant articles are included in the review. Finally, we will review the reference list of included articles to identify any other publications that have not been identified through previous searches.

### Stage 3: Identify Relevant Studies

Identified records will be imported into Covidence. Following the removal of duplicates, two reviewers will independently screen titles and abstracts according to the prespecified eligibility criteria. Any disagreements will be resolved through discussion or referral to a third reviewer.

Following title and abstract screening, full texts will be sourced where possible for the potentially eligible records. Two independent reviewers will then read and screen the full texts against the eligibility criteria, and any disagreements will be resolved through discussion or referral to a third reviewer.

The full screening process will be presented in a PRISMA flow chart. Studies with multiple publications will be grouped and reported as single studies under a primary reference.

### Stage 4: charting the data

#### Data charting process

Data charting forms will be developed to capture all relevant data items related to the review’s aim. Two reviewers will pilot test these forms and carry out the data extraction process.

#### Data Items

To address the review aims, the following data items will be extracted from included articles: author(s), year of publication, country of publication or data collection, aims/purpose, population and sample size within the source of evidence (if applicable), methodology, key findings that relate to the scoping review question/s, curricula activities, interventions, disciplines, and process/strategy used for IP MH identity formation.

### Stage 5: collating, summarizing and reporting the results

#### Synthesis of results

We will carry out a narrative synthesis to present a descriptive overview over included articles. We will use this synthesis to develop a conceptual model for the process of creating an interprofessional field identity in mental health.

## Discussion

Through reviewing the literature, this scoping review will highlight those aspects which are essential to help achieve an interprofessional, mental health, identity in pre-registration students. To date, research on interprofessional identity has focused on an identity across professions, rather than an identity across the mental health field, and particularly at pre-registration level. Through undertaking this review it is anticipated that a theoretical model to help facilitate an interprofessional mental health identity can be proposed.

## Data Availability

No datasets were generated or analysed during the current study. All relevant data from this study will be made available upon study completion.

## Appendix 1 Medline search strategy

1. interprofession*.tw.
2. interdisciplin*.tw.
3. interoccupation*.tw.
4. interinstitut*.tw.
5. interagen*.tw.
6. exp Interdisciplinary Communication/
7. intersector*.tw.
8. interdepartment*.tw.
9. interorgani?ation$.tw.
10. interprofessional relations/
11. multiprofession*.tw.
12. multidisciplin*.tw.
13. multiinstitution*.tw.
14. multioccupation*.tw.
15. multiagenc*.tw.
16. multisector*.tw.
17. multiorgani?ation*.tw.
18. transprofession*.tw.
19. transdisciplin*.tw.
20. collaborat*.tw.
21. 1 or 2 or 3 or 4 or 5 or 6 or 7 or 8 or 9 or 10 or 11 or 12 or 13 or 14 or 15 or 16 or 17 or 18 or 19 or 20
22. (education* or train* or learn* or teach* or course* or curricul*).tw.
23. Education, Nursing, Associate/ or Education, Medical, Undergraduate/ or Nursing Education Research/ or Education, Graduate/ or Education/ or Education, Pharmacy/ or Education, Pharmacy, Continuing/ or Education, Nursing, Baccalaureate/ or Interprofessional Education/ or Education, Professional/ or Education, Public Health Professional/ or Education, Continuing/ or Education, Medical, Continuing/ or Education, Nursing/ or Education, Nursing, Continuing/ or Education, Nursing, Graduate/ or Education, Medical/ or Education, Dental/ or Education, Pharmacy, Graduate/ or Education, Medical, Graduate/ or Education, Dental, Continuing/
24. student*.tw.
25. 22 or 23 or 24
26. (psychiat* or psycho* or mental* or emot*).tw.
27. exp Mental Health/
28. exp Mental Health Services/
29. 26 or 27 or 28
30. 21 and 25 and 29

## References

1. Votruba N, Thornicroft G. Sustainable development goals and mental health: learnings from the contribution of the FundaMentalSDG global initiative. Global Mental Health. 2016;3:e26. Epub 2016/09/09. doi: 10.1017/gmh.2016.20.

2. World Health Organisation. Framework for Action on Interprofessional Education and Collaborative Practice 2010 [cited 2025 7th April]. Available from: https://www.who.int/hrh/resources/framework_action/en/.

3. Reeves S, Pelone F, Harrison R, Goldman J, Zwarenstein M. Interprofessional collaboration to improve professional practice and healthcare outcomes. Cochrane Database of Systematic Reviews. 2017;(6). doi: 10.1002/14651858.CD000072.pub3. PubMed PMID: CD000072.

4. Lapkin S, Levett-Jones T, Gilligan C. A systematic review of the effectiveness of interprofessional education in health professional programs. Nurse Education Today. 2013;33(2):90–102. doi: 10.1016/j.nedt.2011.11.006.

5. Khalili H, Carole O, Spence LHK, and Farah R. An interprofessional socialization framework for developing an interprofessional identity among health professions students. Journal of Interprofessional Care. 2013;27(6):448–53. doi: 10.3109/13561820.2013.804042.

6. Reinders JJ, Lycklama À Nijeholt M, Van Der Schans CP, Krijnen WP. The development and psychometric evaluation of an interprofessional identity measure: Extended Professional Identity Scale (EPIS). J Interprof Care. 2020:1–13. Epub 20200203. doi: 10.1080/13561820.2020.1713064. PubMed PMID: 32013632.

7. Reinders J-J, Krijnen W. Interprofessional identity and motivation towards interprofessional collaboration. Medical Education. 2023;57(11):1068–78. doi: 10.1111/medu.15096.

8. Witz A. Professions and Patriarchy. London: Routledge; 1992.

9. Freidson E. Profession of Medicine. New York: Harper and Row; 1970.

10. Nyatanga L. The Archetypal Roots of Ethnocentrism. In: Colyer H, Helme, M., and Jones, I., editor. The Theory-Practice Relationship in Interprofessional Education. London: Higher Education Academy 2005.

11. Nursing & Midwifery Council. The code: Professional standards of practice and behaviour for nurses, midwives and nursing associates. 2018.

12. Buescher T, and McGugan S. Standing out on the Margins: Using Dialogical Narrative Analysis to Explore Mental Health Student Nurse Identity Construction and Core Modules. Issues in Mental Health Nursing. 2022;43(8):737–47. doi: 10.1080/01612840.2022.2037174.

13. Owens M. An exploration of collaborative practice and non-formal interprofessional education by medical and nursing students in the primary care setting University of Huddersfield; 2011.

14. Price SL, Meaghan S, Victoria L, Joan A, Cynthia A, Harriet D, et al. Pre-entry perceptions of students entering five health professions: implications for interprofessional education and collaboration. Journal of Interprofessional Care. 2021;35(1):83–91. doi: 10.1080/13561820.2019.1702514.

15. Abrams D. Social Identity, Psychology of. In: Smelser NJ, Baltes PB, editors. International Encyclopedia of the Social & Behavioral Sciences. Oxford: Pergamon; 2001. p. 14306–9.

16. Arksey H, and O’Malley L. Scoping studies: towards a methodological framework. International Journal of Social Research Methodology. 2005;8(1):19–32. doi: 10.1080/1364557032000119616.

